# Associations with Prevalent Sexually Transmitted Infection Prevalence among Young Women Domestic Workers in Central Uganda

**DOI:** 10.1101/2025.08.21.25334130

**Authors:** Annet A. Onzia, Andrew Mujugira, Vivian Nakate, Joseph Musaazi, Justine Bukenya, Eric Sseggujja, Patricia A. Ajeru, Rosalind Parkes-Ratanshi, Johan H. Melendez, Matthew M. Hamill, Yukari C. Manabe, Barbara Castelnuovo

**Author notes:** Corresponding author: Annet Onzia Aketoko, Infectious Diseases Institute, College of Health Sciences, Makerere University, P. O. Box 22418, Kampala, Uganda. Shared last authorship (Yukari C. Manabe, Barbara Castelnuovo). Alternate Contact: Barbara Castelnuovo, PhD, Infectious Diseases Institute, College of Health Sciences, Makerere University, P.O. Box 22418, Kampala, Uganda.

## Abstract

**Objective:** Our study aimed to determine the prevalence and correlates of sexually transmitted infections (STIs) among adolescent girls and young women (AGYW) employed as household domestic workers in Kampala, Uganda.

**Methods:** Ugandan AGYW aged 14-24 participated in a community-based cross-sectional study in the Kampala Metropolitan Area from November 2023 to March 2024. Self-collected vaginal swabs were tested for *Neisseria gonorrhoeae* (NG) and *Chlamydia trachomatis* (CT) using GeneXpert™. Testing for HIV and syphilis was performed using the Bioline™ HIV/syphilis Duo test. Correlates of STI prevalence were determined using modified Poisson regression.

**Results:** Of 262 AGYW enrolled, the median age was 20 years (IQR 18-23), and 87 (33%) had an STI: *Chlamydia trachomatis* (27.5%), syphilis (5.7%), HIV (4.6%), and *Neisseria gonorrhoea* (1.5%). Overall, 14.9% had >1 STI. Of 12 AGYW with HIV, three (25%) were newly diagnosed, and 6/9 had detectable viral loads (≥1000 copies/ml). In the prior 12 months, 126 (48%) had received syndromic STI treatment, but only 35 (28%) notified their partners. Additionally, 102 (39%) had used emergency contraception, with 54% believing it was protective against STIs, including HIV. Factors associated with STI prevalence included receipt of non-cash (instead of salary) remuneration for domestic work (adjusted prevalence ratio [aPR] 2.26; 95% CI:1.04-4.92; P=0.040), lower education attainment (aPR 1.73; 95% CI: 1.16-2.57; P=0.007), alcohol consumption in past six months (aPR 1.72; 95% CI: 1.12-2.64; P=0.013) and transactional sex during the past six months (aPR 1.43; 95% CI: 1.01-2.04; P=0.045). Conversely, self-reported sexual abuse was negatively associated with STIs (aPR 0.48; 95% CI: 0.28-0.83; P=0.008).

**Conclusion:** The high prevalence of undiagnosed STIs and unsuppressed HIV among AGYW domestic workers in Kampala highlights the urgent need to increase HIV/STI prevention, testing and treatment coverage for these underserved and vulnerable young women. Targeted interventions are needed to mitigate STI/HIV acquisition in this population.

**What is already known on this topic - summarise the state of scientific knowledge on this subject before you did your study, and why this study needed to be done.:** Adolescent girls and young women (AGYW) in Uganda who are out of school and employed in household domestic work are a highly vulnerable population, often migrating from rural communities to cities with consequent social isolation, and are subject to unequal power dynamics with employers. This puts them at risk of intergenerational and transactional sex. The prevalence of STIs and HIV, and the associated socio-behavioural factors, have been poorly studied in this demographic, limiting the development of interventions for this group.

**What this study adds - summarise what we now know as a result of this study that we did not know before.:** This cross-sectional study, which estimated STI and HIV prevalence among young females employed in household domestic work, found that one-third of AGYW domestic workers were diagnosed with at least one STI. The findings reveal a high burden of STIs and HIV, with many experiencing co-infections. Contributing factors such as poor working conditions, limited educational attainment and prevalent sexual behaviours associated with STIs– including transactional sex, sexual abuse, alcohol use, and inconsistent condom use– exacerbate their vulnerability.

**How this study might affect research, practice, or policy - summarise the implications of this study.:** This research provides insights into STI and HIV risks for vulnerable AGYW employed in domestic work. It highlights the need for tailored HIV and STI interventions that focus on increased awareness, economic and educational empowerment, improved working conditions, socio-behavioural changes, legal redress, as well as expanded access to STI/HIV testing and treatment services for this hard-to-reach population.

## Introduction

Adolescent girls and young women (AGYW) in sub-Saharan Africa (SSA) bear a significant burden of HIV and other sexually transmitted infections (STIs), with about 80% of global HIV acquisitions occurring among AGYW in SSA [1]. Worldwide, more than one million curable STIs are acquired daily, with the prevalence and associated burden relatively higher in SSA [2]. In Uganda, AGYW account for 32% of all new HIV infections, with an estimated 730 infections occurring every week [3]. Data from three consecutive Uganda Demographic Health Surveys show a pooled self-reported STI prevalence of 26.0% among AGYW aged 15-24 years, with 22.0%, 36.3%, and 23.1% reporting an STI infection in the years 2006, 2011, and 2016, respectively [4]. Another study compared STI prevalence among AGYW participating in HIV prevention studies and reported a prevalence of CT at 10.3% and NG at 1.7% in Eastern Africa compared to CT at 15.1% and NG at 4.6% respectively, in South Africa [5]. A study among Ugandan AGYW aged 15-24 years reported NG and/or CT prevalence of 6.8% among those in urban areas compared to 7.8% in rural areas [6]. The elevated rates of HIV and STIs are attributable to biological vulnerabilities, including the immaturity of cervical epithelial tissue, large cervicovaginal mucosal surface area, and prolonged exposure to HIV at mucosal sites [7].

The management of STIs in SSA mainly depends on a syndromic approach due to limited access to etiological testing. This method fails to detect asymptomatic infections, especially in women, leading to many undiagnosed and untreated cases, which in turn fuel ongoing transmission and long-term clinical sequelae [8]. A meta-analysis of STI studies from low- and middle-income countries found that the overall proportions of women with asymptomatic *Chlamydia trachomatis* (CT), *Neisseria gonorrhoeae* (NG), and *Trichomonas vaginalis* (TV) infections were 60.7%, 53.3%, and 56.9%, respectively [9]. These rates were highest among African women with CT, NG, and TV at 68.6%, 67.2%, and 64.9%, respectively [9].

Behavioural vulnerabilities for AGWY include age-disparate relationships that often lead to engagement in high-risk sexual behaviours, such as inconsistent condom usage and transactional sex, compounded by a limited capacity to recognise dangerous situations and adopt protective measures [10]. Furthermore, AGYW face gender-based discrimination and stigma intensified by prevailing societal norms [11], which further increase their vulnerability to HIV and other STIs. Simultaneously, access to healthcare remains limited [10].

Girls in SSA frequently leave school due to cultural traditions like early marriage, gender-based expectations including household responsibilities from an early age, the belief that they do not need as much education as boys, financial hardships like unaffordable school fees and poor menstrual hygiene management [12]. Without access to formal skills training, out-of-school AGYW tend to gravitate towards low-wage jobs, such as domestic work in households. Research indicates that AGYW employed in these low-paying jobs are at an increased risk of acquiring STIs than those who are unemployed, likely due to risky sexual behaviours and limited ability to negotiate safer sex practices [13]. However, little is known about the burden of STIs among the subset of out-of-school girls who are domestic workers. This community-based cross-sectional study aimed to assess the prevalence and correlates of STIs among AGYW employed in the domestic services industry in two districts in Central Uganda.

## METHODS

### Study setting

We conducted a community-based cross-sectional study from November 2023 to March 2024 in six urban and semi-urban communities within the Kampala Metropolitan Area (i.e., Kampala and Wakiso districts), Uganda. Study activities were conducted in the offices of community-based health care workers, locally called Village Health Teams (VHTs), or in designated safe spaces under the Determined, Resilient, Empowered, AIDS-Free, Mentored, and Safe (DREAMS) project, a multi-component intervention designed to address the underlying causes of vulnerability to HIV acquisition among AGYW across SSA [14]. This was located in the local communities of Kasangati Town Council, Nansana division, Nabweru, Wakiso Town Council, and Rubaga and Kawempe divisions in the two districts. The VHTs are non-professional, community-based health volunteers trained by the Ministry of Health to serve as the first point of contact for healthcare at the community level. They promote health, prevent disease, and connect individuals to formal health services.

### Study population

We screened and enrolled AGYW employed as household domestic workers, i.e., those performing household tasks such as cooking, cleaning, laundry, childcare, and gardening. We recruited females aged 14-24 years. Those <18 years had to qualify as emancipated or mature minors, i.e., pregnant, married, had a child or were self-sufficient or employed, according to national research guidelines [15]. We included AGYW who had worked for at least three months in domestic work, had ever had penetrative vaginal sex for ease of vaginal swab self-sample collection, and who were willing to respond to the questionnaire and provide biological samples (vaginal swabs and blood). We collaborated with community health workers and women’s councillors to identify potential participants within their jurisdictional areas, using household registration lists of domestic workers and a door-to-door approach. They briefly explained the purpose of the study and referred the AGYW to the nearest study site, where the research team assessed their eligibility. Trained research nurses obtained written informed consent from eligible participants in either the local language, Luganda or English. We maintained a de-identified enrolment log to track the proportion of persons who declined to enrol and documented reasons for non-participation.

### Data collection and procedures

Two research assistants administered a semi-structured, pre-tested questionnaire, guided by the constructs of the Socio-Ecological model in either Luganda (the local language) or English. Questions were adapted from a similar AGYW study [16]. Before data collection, subject matter experts assessed the questionnaire for validity and reliability. They reviewed the content to confirm it was relevant, appropriate, and comprehensive to ensure the results would be accurate, consistent, and trustworthy. Expert feedback led to questionnaire modifications. We undertook a pilot study with five respondents at one study site to ensure that questions were clear and easily understood. Questions considered ambiguous, unclear, or likely to cause biased responses were removed. The questionnaire included neutral open-ended questions and short-term recall (3-12 months) to reduce social desirability and recall bias regarding sexual behaviours. Participants were assigned anonymous identifiers to protect their identity. Data were recorded on tablet computers using Kobo Toolbox at study sites and backed up daily on a secure, access-restricted server. Automatic range and consistency checks were implemented during data entry, ensuring all questions required a response.

### Sample collection and processing

We provided clear, step-by-step instructions using visual aids and verbally for vaginal swab self-collection to ensure a safe and hygienic process. Research nurses collected venous blood. HIV and syphilis testing were done at the point of care using the Bioline™ HIV/syphilis Duo test (Abbott, Illinois, USA), with STAT-PAK^®^ (Chembio, Medford, NY, USA) and Uni-Gold^TM^ HIV-1/2 (Trinity Biotech, Wicklow, Ireland) used to confirm positive HIV test results, according to the Ugandan testing algorithm guidelines. Clearly labelled samples were transported in cooler boxes to the IDI Translational Laboratory within six hours of collection and tested for NG and CT using the Cepheid GeneXpert^®^ molecular test (Cepheid Europe, HBDC, Maurens-Scopont, France). A reactive Bioline syphilis treponemal antibody was confirmed using the rapid plasma reagin [RPR] (Becton Dickinson-Sparks, Maryland, USA). All participants with positive STI results received standard antimicrobial treatment for curable STIs, and those diagnosed with HIV were linked to antiretroviral therapy (ART) following Uganda guidelines [17]. Those requiring counselling or legal services for sexual and gender-based violence were referred to professional organisations.

### Sample size and statistical analysis

The sample size of 262 AGYW was calculated using Leslie Kish’s formula for cross-sectional prevalence studies. We assumed an expected STI prevalence of 15%, based on findings from a South African study [18]. The calculation was based on a 95% confidence level, a 5% margin of error (precision), and a significance level of α = 0.05. We adjusted the calculated sample size for design effect to account for potential clustering effects, assuming an intra-cluster correlation coefficient (ICC) of 0.0045, given that participants were drawn from six different centres/clusters that could have potentially varying behavioural patterns and risk. The sample size was inflated by 10% to account for potential non-response. A proportionate-to-size allocation strategy was used to determine the number of participants to be recruited from each of the six clusters. Participants were systematically selected within each cluster using a predetermined sampling interval until the required sample size per site was reached.

The primary outcome was the prevalence of laboratory-confirmed CT, NG, syphilis, and HIV, analysed as a composite variable. STI prevalence was defined as the proportion of AGYW who tested positive for any STI. Sexual behaviours that increased vulnerabilities to STI acquisition were defined as any of the following: sexual debut at age <18 years (age of consent under Ugandan law), having >1 sex partner, no condom use at last sex, sometimes/never condom use, emergency contraception use (a proxy measure of condomless sex), alcohol or drug use, transactional sex, and sexual abuse. Sexual abuse was defined as non-consensual vaginal, anal, or oral sex. Chi-Square or Fisher’s exact test were used to compare STI prevalence across participant characteristics. Modified Poisson regression was used to examine factors associated with STIs and estimate adjusted prevalence ratios and 95% confidence intervals. Variables in the unadjusted model with p>0.2 were included in the adjusted model. All statistical analyses were conducted using STATA 17 (College Station, Texas, USA).

### Statement of ethics approval

The study received ethical approval from Makerere University School of Public Health Research Ethics Committee (MaKSPH-REC 262) and administrative clearance from the Uganda National Council for Science and Technology (HS3152ES). Written informed consent was obtained from all participants in Luganda or English, with impartial witnesses used for those deemed illiterate. We enrolled participants aged 15-17 (below the legal age of consent), as recommended by national research guidelines [15]. All participants received USD 5 as reimbursement for their time.

## RESULTS

We screened 306 participants and enrolled 262 (85.6%). Reasons for non-enrolment included being in school (n=14), fear of HIV testing (n=9), failure to complete study activities (n=7), fear of vaginal swab self-collection (n=6), lack of time for study activities (n=4), and being ≥24 years old (n=4). The median age was 20 (interquartile range [IQR]18-23), and 29% were ≤ 18 years old. Most (61%) had attained a primary educational level (≤7 years of schooling), and 5% were married. Nearly all (96%) were formally employed, 84% were live-in domestic workers who were paid in cash, and 73% earned a monthly salary between USD 14 and 27. Most (70%) AGYW reported sexual debut <15 years, while 27% had ≥2 sexual partners in the past six months. Only 28% used condoms during their last sexual encounter, and 9% reported consistent use. Over half (56%) engaged in transactional sex, 39% used emergency contraception, and 11% used alcohol. A significant portion (21%) reported sexual abuse, for which 55% did not seek help due to fear of job loss or threats; employers were the most common perpetrators (63%). Overall, 75% of participants reported ≥2 high-risk behaviours, and nearly 40% had at least four (**Table 1**). Only 16% of participants had been engaged in the DREAMS program.

**Table 1.** Socio-demographic and behavioural characteristics of adolescent girls and young women domestic workers.

| **Variable** | **N (%) or median (IQR)** |
| --- | --- |
| **Age, median (IQR)** | 20 (18-23) |
| ≤18 years | 77 (29.4) |
| >18 years | 185 (70.6) |
| **Marital status** |  |
| Married and monogamous | 13 (5.0) |
| Single with no sexual partner | 106 (49.6) |
| Single with a regular sexual partner | 130 (40.5) |
| Separated/divorced | 13 (5.0) |
| **Highest education level** |  |
| Primary | 159 (60.7) |
| Post primary | 103 (39.3) |
| **Domestic worker status** |  |
| Live in worker | 219 (83.6) |
| Live out (comes and goes everyday) | 43 (16.4) |
| **Renumeration method** |  |
| Cash renumeration | 252 (96.2) |
| Non-cash renumeration | 10 (3.8) |
| **Engagement in DREAMS program** |  |
| Yes | 41 (15.6) |
| No | 221 (84.4) |
| **Monthly pay (USD), n=252** |  |
| <14 | 38 (15.1) |
| 14-27 | 185 (73.4) |
| >27 | 29 (15.1) |
| **Participants known to be living with HIV at study entry (n=9)** |  |
| **ART Treatment** |  |
| Yes | 9 (100) |
| **Viral load status** |  |
| Detectable | 0 |
| Undetectable | 3 (33.3) |
| Unknown | 6 (66.7) |
| **Sex partners in previous 6 months** |  |
| 1 partner | 101 (38.6) |
| ≥2 partners | 70 (26.7) |
| None | 91 (34.7) |
| **Age at sexual debut** |  |
| 8-12 years | 79 (30.1) |
| 13-15 years | 105 (40.1) |
| >15 years | 28 (10.7) |
| Not sure | 50 (19.1) |
| **Frequency of condom use in past 6 months** |  |
| Always | 24 (9.2) |
| Sometimes/rarely | 100 (38.2) |
| Never | 138 (52.7) |
| **Condom use at last sexual encounter** |  |
| Yes | 72 (27.5) |
| No | 190 (72.5) |
| **Transactional sex in past 6 months** |  |
| Yes | 146 (55.7) |
| No | 116 (44.3) |
| **Alcohol use in past 6 months** |  |
| Yes | 29 (11.1) |
| No | 233 (88.9) |
| **Emergency contraception use in past 12 months** |  |
| Yes | 102 (38.9) |
| No | 160 (61.1) |
| Belief that emergency contraception is protective against STIs, *n*=102 |  |
| Yes | 55 (53.9) |
| No | 47 (46.1) |
| **Sexual abuse during employment in domestic work** |  |
| Yes | 56 (21.4) |
| No | 206 (78.6) |
| **Perpetrators of sexual abuse, n=56** |  |
| Employer | 35 (62.5) |
| Relative/friend of employer | 16 (28.6) |
| Another employer in the household/neighbourhood | 2 (3.6) |
| Another person | 3 (5.4) |
| **Sexual abuse reported, n=56** |  |
| Yes | 5 (8.9) |
| No | 51 (91.1) |
| **Reasons for not reporting abuse, n=51** |  |
| I was afraid to lose my job | 28 (54.9) |
| I was threatened to not report | 22 (43.1) |
| I reported a prior experience and was not supported | 1 (2.0) |
| Declined to respond | 1 (2.0) |
| **Risky sexual behaviours** |  |
| ≤2 characteristics | 66 (25.2) |
| >2 characteristics | 196 (74.8) |
| **Risky sexual behaviour** | 3 (2-4) |
| **Sexual behaviours associated with STI acquisition (count of characteristics) \*** |  |
| 1 | 14 (5.3) |
| 2 | 52 (19.9) |
| 3 | 92 (35.1) |
| ≥4 | 104 (39.7) |

*\*Any of the following characteristics: Sexual debut <18 years, >one sexual partner, no condom use at last sexual encounter, emergency contraception use, alcohol use, drug use, transactional sex, and sexual abuse.*
*IQR-interquartile range; DREAMS- Determined, Resilient, Empowered, AIDS-free, Mentored and Safe; HIV-Human Immunodeficiency Virus; ART-Antiretroviral treatment; STIs-sexually transmitted infections*

### STI prevalence

In total, 87 AGYW (33%) had at least one STI, including HIV. STI prevalence was as follows: CT at 27.5% (72/262), syphilis at 5.7% (15/262), with 60% (9/15) having RPR titres ≥1:8, HIV at 4.6% (12/262), and NG at 1.5% (4/262) (**Figure 1**). Among those diagnosed with an STI, 14.9% (13/87) had more than one infection; specifically, ten AGYW had two STIs, while three had three STIs. Of the ten participants with dual STIs, four presented with syphilis and chlamydia, two had chlamydia and gonorrhoea, three had HIV and chlamydia, and one had HIV and syphilis. Among the three individuals who tested positive for three STIs, two had HIV, syphilis, and chlamydia; the remaining participant had chlamydia, syphilis and gonorrhoea. All nine participants known to be living with HIV reported being on ART. However, only one-third (33.3%, n=3) indicated that their last test results showed an undetectable viral load; the remaining six did not know their most recent viral load result. Notably, 25% (3/12) of those living with HIV were unaware of their diagnosis before study participation.

**Figure 1.**
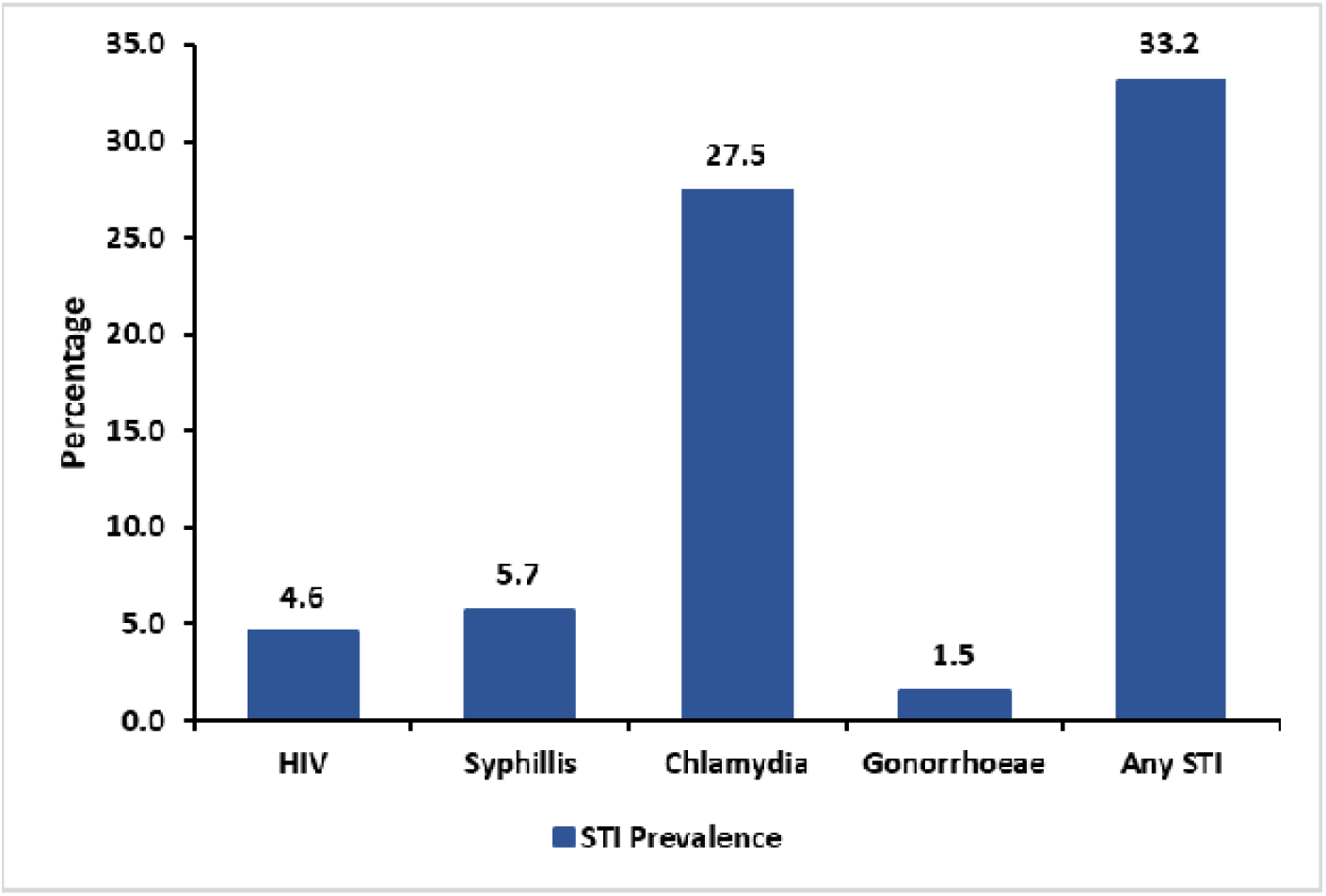
Prevalence of STIs among adolescent girls and young women employed in domestic work

### Factors associated with selected STIs and HIV

Receipt of non-cash remuneration (i.e., food, clothing and lodging instead of salary) (adjusted prevalence ratio [aPR] 2.26; 95% CI:1.04-4.92; P=0.040), lower educational attainment (aPR 1.73; 95% CI: 1.16-2.57; P=0.007), alcohol consumption in past six months (aPR 1.72; 95% CI: 1.12-2.64; P=0.013) and engagement in transactional sex during the past six months (aPR 1.43; 95% CI: 1.01-2.04; P=0.045) were positively associated with STI prevalence. In contrast, reported sexual abuse within the past 12 months was negatively associated with STIs (aPR 0.48; 95% CI: 0.28-0.83; P=0.008) (**Table 2**).

**Table 2.** Factors associated with STIs among adolescent girls and young women employed in domestic work.

| **Variable** | **STI**<br>**Prevalence**<br>**n/N (%)** | **Unadjusted PR**<br>**(95%CI)** | **P-**<br>**value** | **Adjusted**<br>**(95%CI)** | **P-**<br>**value** |
| --- | --- | --- | --- | --- | --- |
| **Overall** | **87/262**<br>**(33.2)** | **N/A** |  | **N/A** |  |
| **Age** |  |  |  |  |  |
| ≤18 years | 23/77<br>(29.9) | *Ref* |  | - | - |
| >18 years | 64/185<br>(34.6) | 1.16 (0.78-1.72) | 0.47 | - | - |
| **Education level** |  |  |  |  |  |
| Post primary | 25/103<br>(24.3) | *Ref* |  | *Ref* |  |
| Primary and below | 62/159<br>(39.0) | 1.61 (1.08-2.38) | 0.018 | 1.73 (1.16-2.57) | **0.007** |
| **Renumeration** |  |  |  |  |  |
| Cash renumeration | 82/252<br>(32.5) | *Ref* |  | *Ref* |  |
| Non-cash<br>renumeration | 5/10 (50.0) | 1.54 (0.80-2.93) | 0.19 | 2.26 (1.04-4.92) | **0.040** |
| **Age at sexual**<br>**debut** |  |  |  |  |  |
| ≤12 years | 26/79<br>(32.9) | 0.97 (0.59-1.59) | 0.90 |  |  |
| 13-15 years | 39/105<br>(37.1) | 1.09 (0.69-1.73) | 0.71 | - | - |
| >15 years | 5/28 (17.9) | 0.53 (0.22-1.27) | 0.15 | - | - |
| Not sure | 17/50<br>(34.0) | *Ref* |  | - | - |
| **Condom use at**<br>**last sex** |  |  |  |  |  |
| Yes | 19/72<br>(26.4) | 0.74 (0.48-1.13) | 0.166 | 0.70 (0.45-1.06) | 0.094 |
| No | 68/190<br>(35.8) | *Ref* |  | *Ref* |  |
| **Transactional sex in past 6 months** |  |  |  |  |  |
| Yes | 56/146<br>(38.4) | 1.44 (1.00-2.07) | 0.053 | 1.43 (1.01-2.04) | **0.045** |
| No | 31/116<br>(26.7) | *Ref* |  | *Ref* |  |
| **Alcohol use in the last 6 months** |  |  |  |  |  |
| Yes | 16/29<br>(55.2) | 1.81 (1.24-2.65) | 0.002 | 1.72 (1.12-2.64) | **0.013** |
| No | 71/233<br>(30.5) | *Ref* |  | *Ref* |  |
| **Sexual abuse in past 12 months as domestic worker** |  |  |  |  |  |
| Yes | 11/56<br>(19.6) | 0.53 (0.30-0.93) | 0.027 | 0.48 (0.28-0.83) | **0.008** |
| No | 76/206<br>(36.9) | *Ref* |  | *Ref* |  |

*Participants with a confirmatory laboratory result of STIs, i.e., HIV, syphilis, chlamydia, or gonorrhoea, were considered as having an STI. PR denotes prevalence ratios; CI, confidence interval; Ref is the reference category for a covariate in the modified Poisson model fitted on 262 complete cases. Variables for the multivariate model were selected using a 0.2 P-value cut-off in the unadjusted analysis of each covariate. None of the variables in the adjusted model had missing values.*
*STIs-sexually transmitted infections, PR-prevalence ratio; n-number of participants with at least one STI; N-total study sample size*

## DISCUSSION

This cross-sectional study found that one-third of AGYW domestic workers were diagnosed with at least one STI, and 15% of them were diagnosed with more than one STI. Moreover, our findings indicate an HIV prevalence of 4.6%, with 25% previously undiagnosed. This is twice the 2.2% prevalence reported by the Uganda AIDS Commission, among the general population of AGYW in Uganda [3]. The overall prevalence of STIs and sexual behaviour that increase risks for STIs among AGYW domestic workers aligns with other key and priority populations in Uganda, including sex workers [19] and AGYW participating in an HIV pre-exposure prophylaxis trial [20].

The vulnerabilities faced by these AGYW may be exacerbated by power imbalances between employers and employees, which contribute to economic exploitation, social isolation, and limited access to healthcare or legal redress [13]. Previous studies show that AGYW frequently change households, seeking better working conditions or engage in high-risk sexual behaviours, increasing their vulnerability [13], but also potentially contributing to STI transmission. This notable STI prevalence among AGYW underscores the need to prioritise STI control in public health initiatives in this population as a fundamental step to achieving epidemic HIV and STI control in this important but underserved demographic.

We observed prevalent sexual behaviours compounded by known factors such as low educational attainment [21] and unstable employment conditions, influenced by existing power dynamics in domestic work, similar to previous research [13]. Education is known to be associated with better sexual and reproductive health outcomes, with female adolescents who are out of school being at higher risk of acquiring HIV and other STIs [22]. Ensuring girls remain in school for as long as possible could reduce their risk of STI acquisition.

Young women who received non-cash compensation, such as food, clothing, and accommodation, and those involved in transactional sex had a higher STI prevalence than those paid in cash, consistent with previous findings [23, 24]. Mirroring other studies, we also noted an association between alcohol consumption and STI diagnosis [25]. This underscores the importance of improving social support, promoting economic empowerment, and improving employment conditions as effective strategies for preventing STIs and HIV in this group of young women. Previous research shows that the DREAMS program resulted in a reduction of HIV incidence and STI symptoms among AGYW across SSA [26]. Given that only 16% of our participants had been engaged in the DREAMS program before participating in our research, we recommend innovative ways to expand its initiatives, such as income-generating activities, substance use prevention including alcohol control measures, peer support, safe spaces, and economic empowerment for this vulnerable population, which may not easily access these services that have been beneficial in previous studies, as part of the broader HIV prevention efforts for AGYW in SSA [26, 27].

Our research revealed a negative association between reported sexual abuse in the past year and STI prevalence, which differs from findings of earlier studies [28]. The underlying cause of this observation remains uncertain. We hypothesise that it could be linked to unmeasured residual confounding, e.g., condom use by perpetrators to avoid unwanted pregnancy, survivors receiving post-rape care, or avoidance of sexual activity stemming from prior sexual trauma.

The strengths of our study include the proportionate to size and systematic sampling strategies contributed to a robust study design and methodology, yielding valid results that may be generalisable to other AGYW domestic workers in this setting. Additionally, we conducted reference laboratory STI testing, which enabled us to correlate self-reported sexual behaviours with etiologic STI diagnosis. The limitations of this cross-sectional study include the inability to determine temporal relationships, thus limiting our analysis of behavioural correlates associated with STI diagnoses. We did not assess STI symptoms and how they correlate to STI positivity, nor did we test for some STIs which are likely to be common in this population, e.g., *Trichomonas vaginalis*. Our study was susceptible to recall bias and relied on self-reported data for sexual behaviour. This could have introduced response and social desirability biases, although we attempted to minimise this by having same-sex, same-age-group interviewers.

### Conclusions

Our research with Ugandan AGYW employed in domestic work revealed a high prevalence of STIs, often with multiple co-infections. These results highlight the urgent need to strengthen initiatives aimed at keeping girls in school, improving their working conditions, and providing expanded access to STI/HIV testing and treatment. Prioritising this vulnerable group is essential to curtail further transmission and acquisition of STIs.

## Declarations

### Conflict of interest

The authors declare that they have no conflict of interest.

### Author contributions

OAA: Study design, writing original protocol draft, literature search, project administration, data collection, supervision, methodology, manuscript drafts. AM: Data interpretation, draft manuscript writing, and manuscript reviews. VN: Data management, data analysis, data interpretation, manuscript reviews. JM: Methodology, statistical analysis, manuscript review. JB: Study design, protocol and questionnaire draft reviews, supervision, manuscript reviews. ES: Study design, protocol and questionnaire draft reviews, supervision, manuscript reviews. PA: Project administration, data collection, data management, manuscript reviews. RPR: Supervision, study design, manuscript reviews. JHM: Study design, supervision, manuscript reviews. MMH: Study design, supervision, manuscript reviews. YCM: Study design, supervision, funding acquisition, draft manuscript writing, manuscript reviews. BC: Study design, supervision, funding acquisition, draft manuscript writing, and reviews.

### Funding

Research reported in this publication was supported by the Fogarty International Centre of the National Institutes of Health under award number D43TW009771 “HIV and co-infections” (MPI: Kambugu, Castelnuovo, Manabe) and partially through the National Institute of Biomedical Imaging and Bioengineering of the National Institutes of Health under Award Number U54EB007958 (YCM). The content is solely the responsibility of the authors and does not necessarily represent the official views of the National Institutes of Health.

### Data availability statement

The data supporting the findings of this study are available from the corresponding author upon reasonable request. The dataset has been de-identified to protect participant confidentiality and is not publicly available due to ethical and privacy restrictions imposed by the Institutional Review Board and the Uganda National Council of Science and Technology. Researchers who meet the criteria for access to confidential data may request access by contacting the corresponding author, Annet A. Onzia, at.

## Acknowledgements

The authors thank the study participants, data collectors, the local council leaders, and the community healthcare workers at each study site for their contributions to this research.

## Notes

### Competing Interest Statement

The authors have declared no competing interest.

### Author Declarations

Ethics committee of Makerere University School of public health (MaKSPH-REC-262) gave ethical approval for this work

